# Contribution of *de novo* retroelements to birth defects and childhood cancers

**DOI:** 10.1101/2024.04.15.24305733

**Authors:** Chong Chu, Viktor Ljungström, Antuan Tran, Hu Jin, Peter J. Park

**Affiliations:** Department of Biomedical Informatics, Harvard Medical School, Boston, MA, USA

## Abstract

Insertion of active retroelements—L1s, *Alu*s, and SVAs—can disrupt proper genome function and lead to various disorders including cancer. However, the role of *de novo* retroelements (DNRTs) in birth defects and childhood cancers has not been well characterized due to the lack of adequate data and efficient computational tools. Here, we examine whole-genome sequencing data of 3,244 trios from 12 birth defect and childhood cancer cohorts in the Gabriella Miller Kids First Pediatric Research Program. Using an improved version of our tool xTea (x-Transposable element analyzer) that incorporates a deep-learning module, we identified 162 DNRTs, as well as 2 pseudogene insertions. Several variants are likely to be causal, such as a *de novo Alu* insertion that led to the ablation of a whole exon in the *NF1* gene in a proband with brain tumor. We observe a high *de novo* SVA insertion burden in both high-intolerance loss-of-function genes and exons as well as more frequent *de novo Alu* insertions of paternal origin. We also identify potential mosaic DNRTs from embryonic stages. Our study reveals the important roles of DNRTs in causing birth defects and predisposition to childhood cancers.

## INTRODUCTION

Three types of retroelements are still active in the human genome: long interspersed element-1 (L1), *Alu*, and SINE-VNTR-*Alu* (SVA). These retroelements replicate through RNA intermediates by a “copy and paste” mechanism mediated by the LINE-1-encoded proteins; the L1 machinery can also mediate retroduplication of protein-coding genes to generate processed pseudogenes (PPGs). Retroelement insertions into genes may disrupt the function of the gene, potentially leading to a wide spectrum of diseases (Hancks and Kazazian 2016; Burns 2017; Chuong et al. 2017). In particular, *de novo* retroelement (DNRT) insertions have been associated with several developmental disorders and other genetic diseases (Gardner et al. 2019; Brandler et al. 2016; Brandler et al. 2018; Werling et al. 2018; Borges-Monroy et al. 2021). Such DNRT insertions may occur in the gonadal tissues, resulting in a heterozygous germline variant in the proband, or during embryonic development, resulting in a mosaic variant in the proband.

Compared to other types of *de novo* mutations, DNRTs have been less well studied due to *(i)* a lack of large whole-genome sequencing (WGS) datasets (especially trios, which are critical for accurately finding *de novo* insertions), and (*ii*) a lack of reliable computational methods designed specifically for DNRTs. Although several tools have been developed for identifying germline (Gardner et al. 2017; Keane et al. 2013; Thung et al. 2014; Zhuang et al. 2014) and somatic (Lee et al. 2012; Tubio et al. 2014) retroelement insertions, they typically give many high false positive calls for DNRTs because of the low rate of DNRTs and the low variant allele frequency (VAF) of mosaic events. For trio data, one could treat a trio as two pairs of case-control samples and find events that exist in the proband but are absent in both parents; however, we find that a more sensitive detection requires more sophisticated filtering steps specifically designed for the trio design.

Here, we extend our xTea (Chu et al. 2021) pipeline for *de novo* retroelement insertion identification by further integrating a newly developed machine learning based filtering module. We apply our pipeline to 3,244 trios from the Gabriella Miller Kids First Pediatric Research Program (GMKF) composed of 12 cohorts of different birth defects and childhood cancers, as well as 596 trios from the 1000 Genomes Project (Byrska-Bishop et al. 2022) as reference (Tab. S1). We identified 162 DNRTs from the GMKF cohorts, several of which mobilized to genes where disruptive variants previously have been deemed causative for the diseases. Below, we describe several analyses including detailed examination of likely pathogenic insertions, trio-based phasing to determine whether paternal and maternal contributions are equal, identification of mosaic insertions that occurred at early embryonic stages and PPG insertions, characterization of genes with a higher burden of insertions, and the activity of different SVA subfamilies.

## RESULTS

### *De novo* retroelements identification in Kids First and 1KGP data

We built an efficient pipeline for DNRT identification (Fig. 1a). Given trio data, we first run the xTea germline module on the proband to identify a set of initial candidates. Because many of the *de novo* events are potentially mosaic events in the proband with low VAFs, we set lenient criteria including on the VAF threshold to ensure high sensitivity. Next, we run the xTea somatic module on the initial candidates with each of the parents as a control. The output from this step still has a high fraction of false positives due to the low cutoff settings. Thus, we apply two additional filtering steps. First, we convert the *de novo* insertion identification problem to an image classification problem by training a machine learning model based on training data labeled from both real and simulated data (see Methods for details). Second, we manually inspect the candidates to curate the final set of DNRTs. Combining the low cutoffs with the filtering steps allows us to design a pipeline with both high sensitivity and high specificity, while its efficient implementation enables application on large cohorts.

**Fig. 1:**
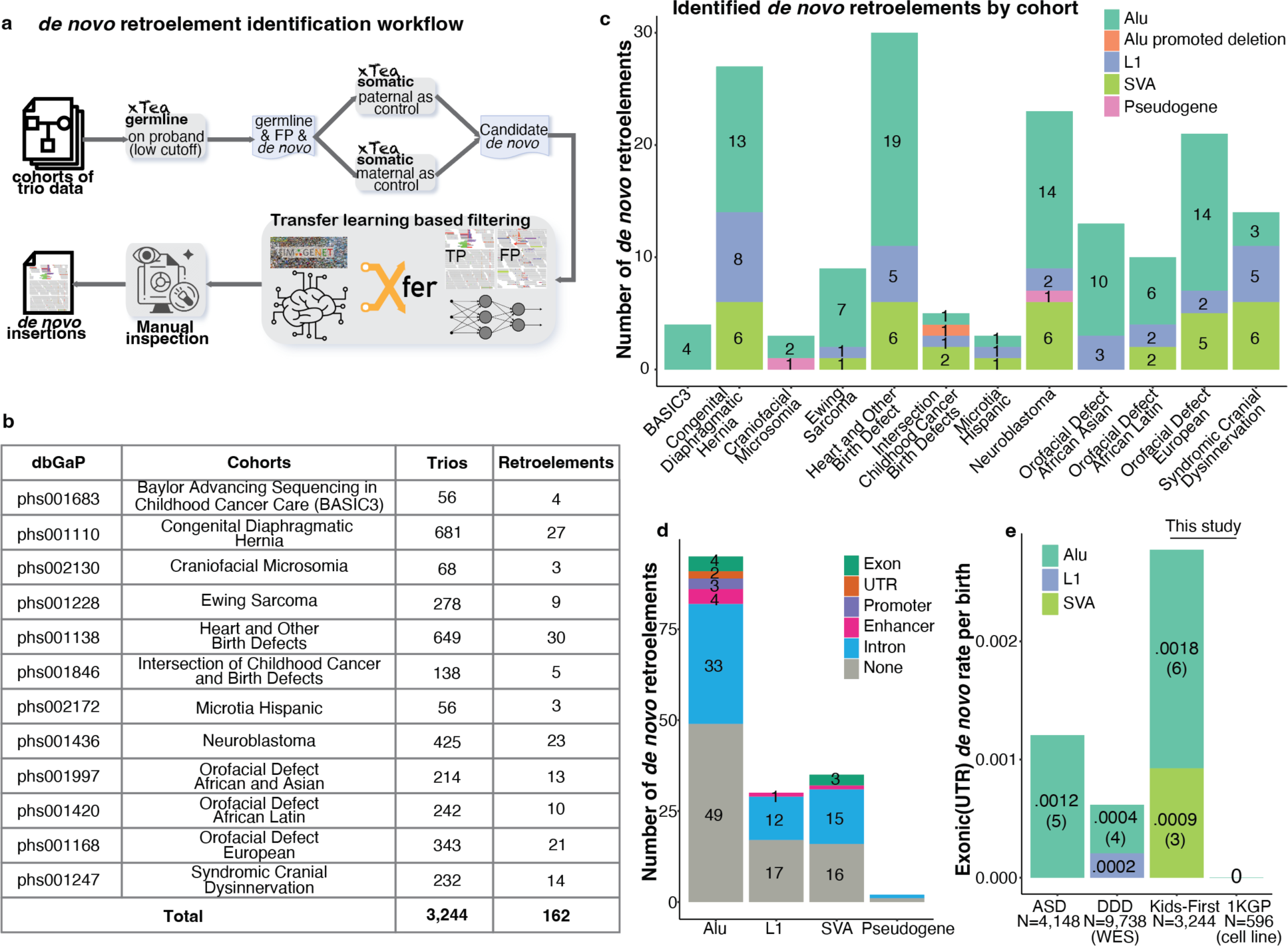
Overview of the pipeline and identified *de novo* retroelements. **a** Schematic outline of the upgraded xTea workflow. The proband sample from trio data is analyzed using the germline module with low cutoffs and subsequently filtered using the somatic module and incorporating parental data to identify candidate *de novo* events. A transfer learning based filtering step following a manual inspection step is applied to filter out the false positives. **b** We ran our pipeline on 3,244 trios from 12 disease cohorts released by the GMKF studies and identified 162 *de novo* retroelements. **c** The number of identified *de novo* retroelements by cohorts and repeat types. Besides the classical TE insertions, we also identified 1 *Alu* promoted deletion and 2 pseudogene insertions from 3 different cohorts. **d** 7 (4 *Alu* and 3 SVA), 2 (*Alu*), 3 (*Alu*), and 6 (4 *Alu*, 1 L1 and 1 SVA) *de novo* retroelements fell in exons, UTRs, promoters, and enhancers respectively. Out of the rest, 33 *Alu*, 12 L1, 15 SVA and 1 pseudogene are intronic insertions, and the remaining are all intergenic ones. **e** The exonic/UTR *de novo* rate from the GMKF and 1KGP cohorts (this study) compared to the Deciphering Developmental Disorders (DDD) study and a study of autism spectrum disorders (ASD).

We ran our pipeline on 12 GMKF cohorts (Fig. 1b), totaling 3,244 WGS trios (Tab. S1). Across all cohorts, we identified 162 DNRTs, including 95 *Alu*, 30 L1, 35 SVA, and 2 PPG insertions (Tab. S2). Fig. 1c shows the number of DNRTs identified from each cohort by repeat type. Besides the classic *Alu*, L1, and SVA insertions, we also identified 1 *Alu*-promoted deletion and 2 PPG insertions from 3 different cohorts. Of the 162 *de novo* insertions, 7 (4 *Alu* and 3 SVA) are exonic, 2 *Alu* affect UTRs, 3 *Alu* are within promoter regions, and 6 (4 *Alu*, 1 L1, and 1 SVA) fall in enhancer regions. Among the others, 33 *Alu*, 12 L1 and 15 SVA are intronic and the rest are intergenic (Fig. 1d). We also analyzed the recently-released trio data from the 1000 Genomes Project (1KGP) consisting of 603 trios of which 596 were successfully processed, leading to identification of 26 *Alu*, 12 L1, and 8 SVA DNRTs (Tab. S3).

We identified 9 and 0 exonic/UTR DNRTs from the GMKF (3,244 births) and 1KGP (596 births) cohorts, respectively. In comparison, the Deciphering Developmental Disorders (DDD) study (Gardner et al. 2019) revealed 6 exonic/UTR variants in 9,738 births analyzed with whole-exome sequencing (WES); another study (Borges-Monroy et al. 2021) of autism spectrum disorder (ASD) showed 5 exonic/UTR variants in 4,184 births (Tab. S4). Thus, the exonic (including UTR) *de novo* rate from the GMKF cohorts in this study is 4.5× (0.0027 vs 0.0006) and 2.25× (0.0027 vs 0.0012) higher than in the DDD and ASD studies, respectively. An earlier systematical study on DDD (Deciphering Developmental Disorders Study, 2017) has shown that developmental disorders have higher *de novo* mutation rate than autism, which is concordant with the much higher rate in the GMKF study than in the ASD study. The much lower rate of the DDD study may be due to the difference in the cohorts or possibly due to the lower sensitivity of the method they used.

### Potentially pathogenic *de novo* retroelement insertions

WGS data on trios provide an opportunity to identify pathogenic mutations and discover novel disease-associated genes. We prioritized the identified 162 DNRTs by checking whether they occur in *(i)* exonic or UTR regions; *(ii)* promoter or enhancer regions, as annotated in ANNOVAR (Yang and Wang 2015); (*iii*) genes that are associated with the disease. In addition, to account for potential compound heterozygosity, we also checked whether there are detectable second hits for those DNRTs that fall in autosomal recessive genes. In Table 1, we show the selected exonic, UTR, and other potentially pathogenic DNRTs and their annotations.

**Table 1.**
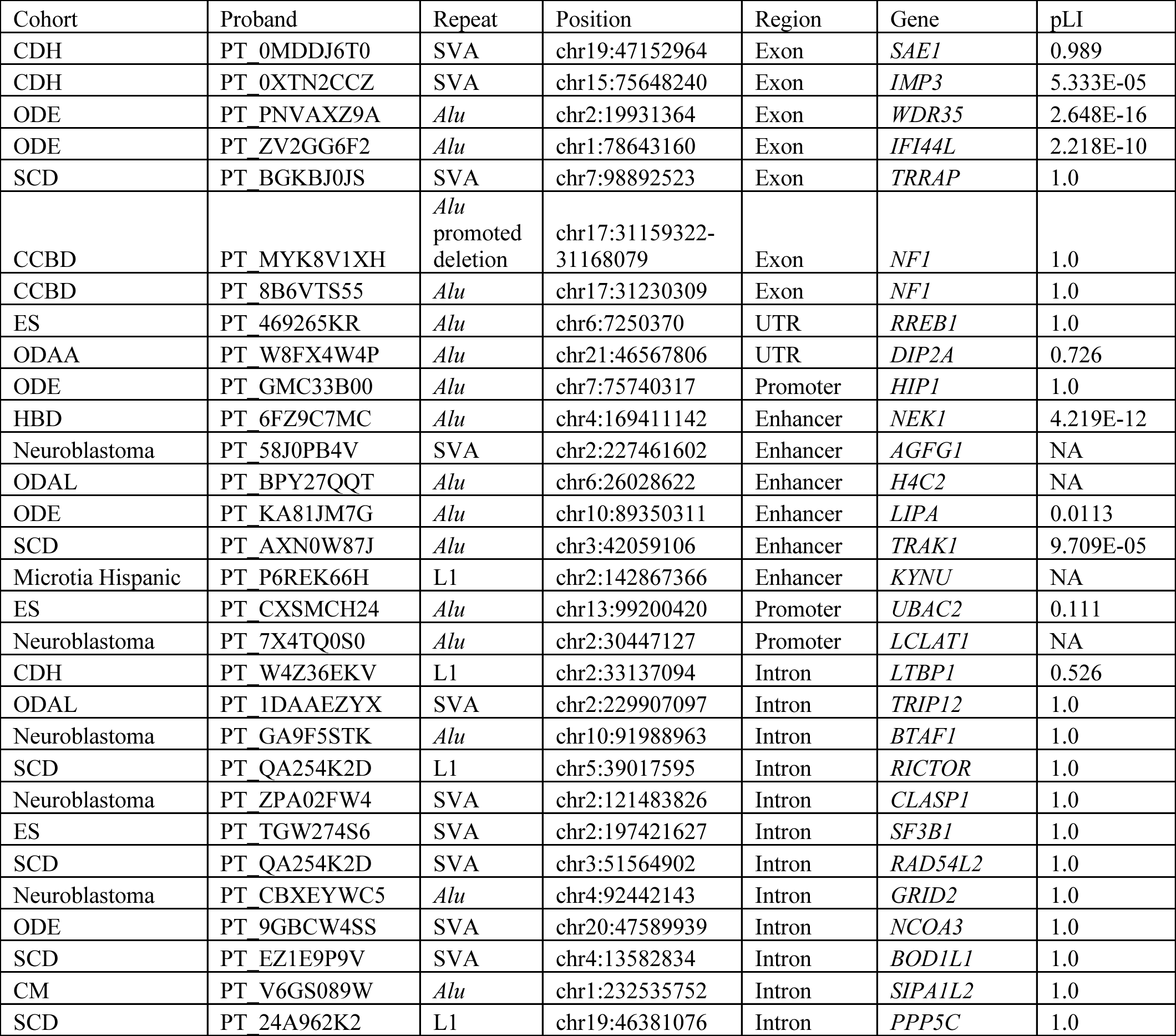
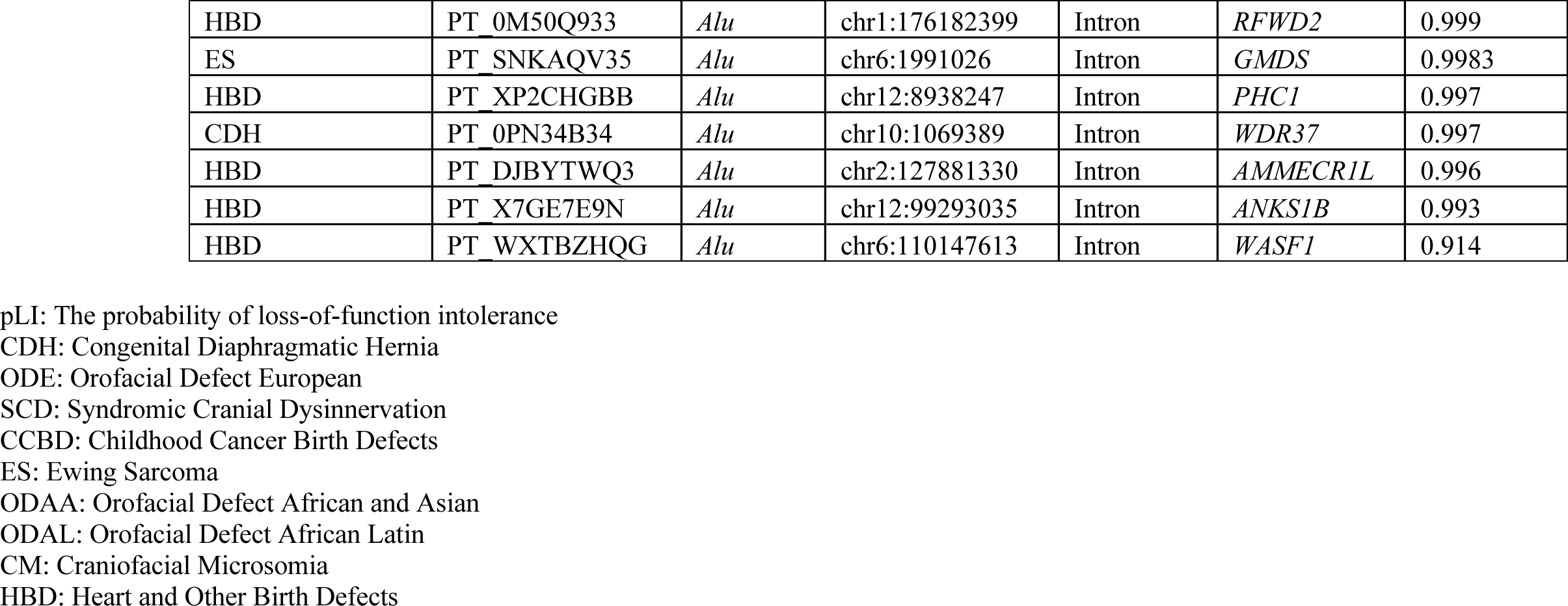
Potential pathogenic *de novo* retroelement insertions identified from the GMKF cohorts.

We identified two *Alu* insertions that fall in the exonic regions of *NF1*. One insertion promoted an 8,758 bp deletion (chr17:31159322-31168079 on hg38) spanning all of exon 4 (Fig. 2a). The polyA reads at the breakpoints, discordant pairs, and the copy number change strongly support the existence of this complex event. Trio-based phasing and the high VAF indicates this complex event is inherited from the father (a mosaic mutation in the father), but we cannot rule out the possibility that this is an early embryonic event (a mosaic mutation in the child). The other *Alu* insertion was mobilized to the 24th exon of *NF1*. Fig. 2b shows the clipped reads, polyA tail, and the discordant reads present in the proband but absent in the parents. Both probands presented with brain tumor and, since neurofibromatosis type 1 caused by a mutation in *NF1* is an autosomal dominant disorder associated with brain tumor, we propose these two exonic *de novo Alu* insertions to be causal mutations. Besides the *NF1* cases, we also identified one SVA insertion mobilized into an exon of the *TRRAP* gene in one proband who was diagnosed with Ptosis (HP:0000508) within the Syndromic Cranial Dysinnervation (SCD) cohort. There have been several reports demonstrating that variants in *TRRAP* are causative for several developmental diseases including autism and syndromic intellectual disability (Cogné et al. 2019; Xia et al. 2019; Mavros et al. 2018), although no directly related cases have been reported for SCD. In addition, we also identified one *de novo Alu* insertion in the 3’ UTR region of *RREB1* in one proband with Ewing Sarcoma. The *RREB1* gene has been associated with Ewing Sarcoma in several of the earlier studies (Shi et al. 2020; Machiela et al. 2018).

**Fig. 2:**
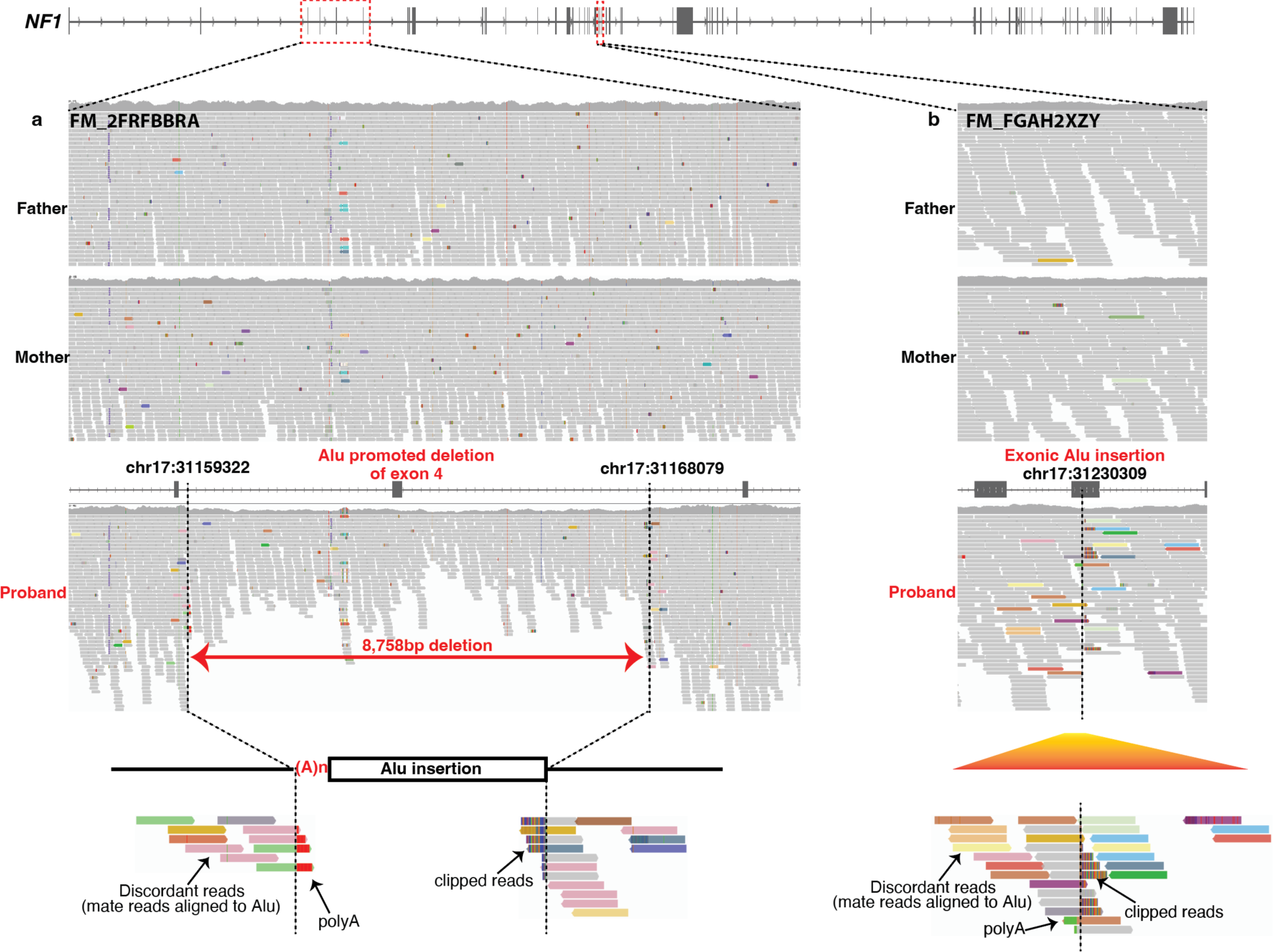
Two exonic *de novo* Alu insertions identified on *NF1* gene. **a** An *Alu* insertion promoted deletion was identified in one trio. The deletion spans 8,758bp and the two breakpoints fell in intron 3 and 4 leading to deletion of the exon of number 4 of *NF1*. The copy number change in the proband shows the existence of the deletion. The clipped reads, polyA reads, and the discordant reads indicate the existence of an *Alu* insertion. Together, all these features demonstrate the existence of the *Alu* insertion promoted deletion. **b** In another trio, one *NF1* exonic *Alu* insertion was identified in proband but absent from parents. Similarly, the clipped reads, polyA reads and discordant pairs strongly support the presence of an *Alu* insertion.

Although it is more challenging to functionally annotate the intronic DNRTs, we identified two cases with variants that have a high chance of explaining the phenotype observed in the proband. One case is a full length *de novo* L1 insertion mobilized in the sense orientation to the intron of *LTBP1.* The patient was diagnosed with congenital diaphragmatic hernia (CDH) (MONDO:0005711). The *LTBP* gene family has been demonstrated to be highly associated with cutis laxa (a connective tissue disorder; Latin for loose skin) in several studies and the disorder is linked to an increased risk of CDH (Bultmann-Mellin et al. 2015; Zhang et al. 2020; Pottie et al. 2021; Urban and Davis 2014). Since most types of cutis laxa are autosomal recessive, we further screened for other types of mutations in the gene and detected one exonic deleterious SNP (2-33567971-C-T; hg38). The SNP is a rare mutation with a population allelic frequency of ∼0.78% in the gnomAD database (Chen et al. 2022). We therefore infer that the *de novo* L1 insertion and the SNV lead to a compound heterozygous state resulting in CDH, although functional studies are needed for validation. The other case is a *de novo Alu* insertion identified in a patient diagnosed with right aortic arch (MONDO:0020417). The insertion mobilized to a strong intronic enhancer of the gene *NEK1*. An on-going study (manuscript in preparation) shows that variants in *NEK1* are associated with congenital heart defect (CHD) in an autosomal dominant pattern.

### Mosaic and parental origin of *de novo* retroelements

Although *de novo* retroelement insertions refer to those identified in the proband but not in the parents, two types of retroelement insertions are identified as *de novo* in practice. One type is a mosaic insertion in a parent that appears in the child as a germline insertion. The other type is a mosaic insertion that occurs in the early development of a proband, with VAF that depends on the timing of the event. As large cohort studies usually have blood or saliva samples with standard depth (∼30X) WGS, only very early embryonic mosaic mutations whose prevalence across tissues is high enough are identified. Earlier trio-based studies on SNVs with ∼30-40X WGS have shown that besides the ∼100 germline *de novo* small mutations (Jónsson et al. 2017; Kong et al. 2012), a small number of proband mosaic mutations were identified (Byrska-Bishop et al. 2022; Ng et al. 2021). Similarly for *de novo* retroelements, we identified variants of both germline and proband mosaic origin.

VAF calculation for retroelement insertions is more challenging compared to SNVs/indels because bias will be introduced when aligning the reads containing repetitive sequences (Fig. S1). We optimized our procedure for retroelements VAF estimation (see Method for details) and calculated the VAF for each of the identified 162, 46, and 144 DNRTs in the GMKF, 1KGP, and ASD cohorts, respectively (Fig. 3a). Note that the VAFs previously reported for the ASD DNRTs (separated by probands and siblings) (Borges-Monroy et al. 2021) used an earlier version of xTea, and they have now been recalculated with the latest version of xTea.

**Fig. 3:**
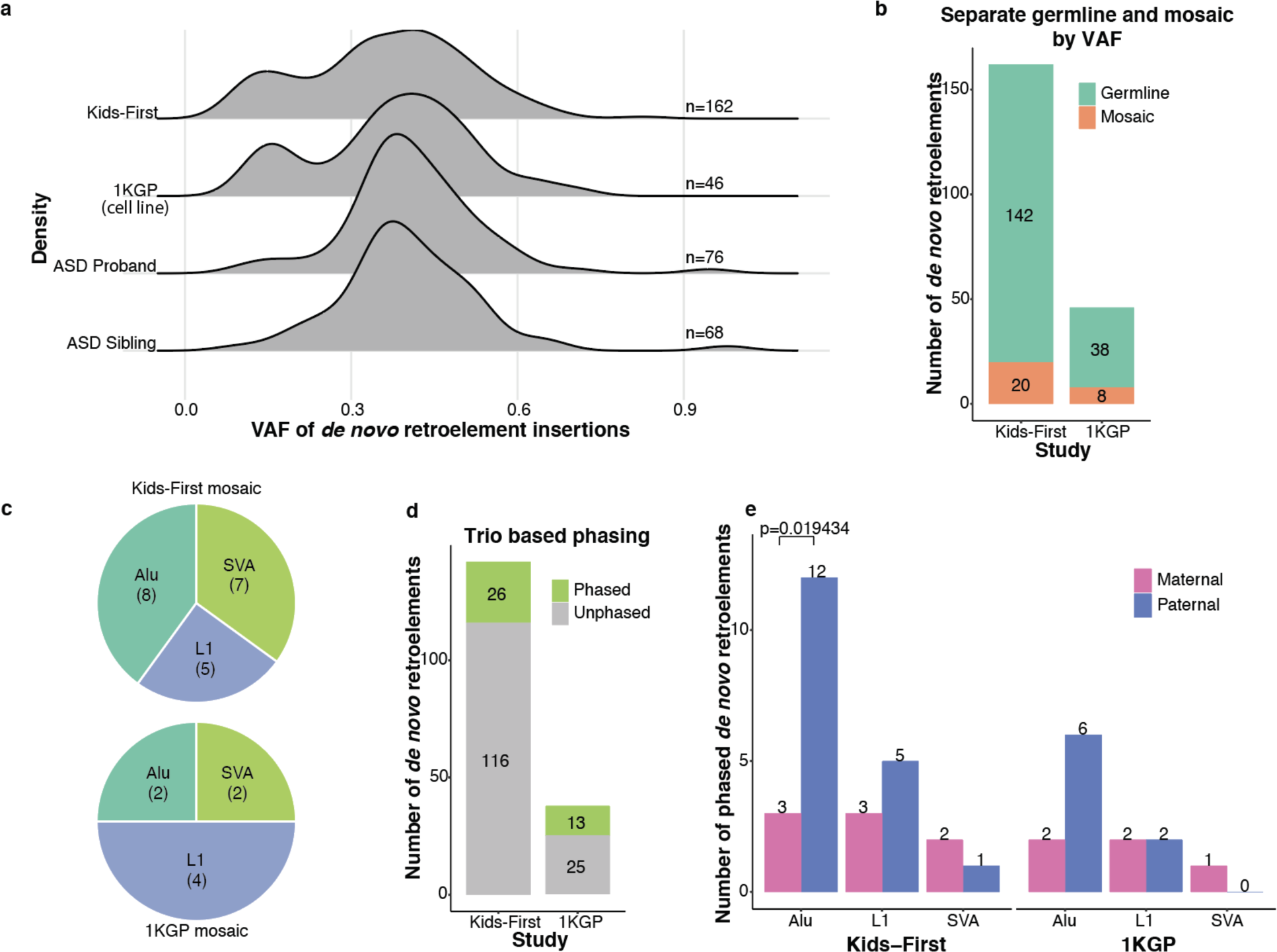
VAF characterization and trio-based phasing of *de novo* retroelements. **a** The variation allele frequency (VAF) for the *de novo* retroelements identified in this study (162 from GMKF and 46 from 1KGP), as well as the 144 (76 from proband and 68 from sibling) reported *de novo* retroelements from the ASD study (Borges-Monroy et al. 2021). The two peaks in density plots of GMKF and 1KGP cohorts indicate the presence of mosaic retroelements in proband. **b** Gaussian mixture model separation of germline and mosaic *de novo* retroelements based on the density of VAF. Out of the 162 and 46 *de novo* retroelements, 20 and 8 mosaic ones were identified for the GMKF and 1KGP respectively. **c** We checked the repeat types of the mosaic events. 8 *Alu*, 5 L1, and 7 SVA were annotated for the 20 GMKF mosaic retroelements, and 2 *Alu*, 4 L1, and 2 SVA were annotated for the 1KGP mosaic ones. **d** For the germline *de novo* retroelements, we applied a trio-based phasing step and identified 26 (out of 142) and 13 (out of 38) phasable ones for the GMKF and 1KGP respectively. **e** Further checking the phased ones from the GMKF cohorts, we found more *de novo Alu* insertions were transmitted from father than from mother (p-value=0.019, exact binomial test), while for L1 and SVA it is unclear. The same pattern was observed from the 1KGP data, although not statistically significant due to the sample size being small.

In the VAF density plot (Fig. 3a), we observed two peaks for GMKF and 1KGP, indicating the existence of germline and mosaic DNRTs in the proband. Using a gaussian mixture model to separate the two event classes, we identified 20 (out of 162) and 8 (out of 46) mosaic DNRTs for the GMKF and 1KGP cohorts, respectively (Fig. 3b). We found mosaic DNRTs for all three types of TE insertions (8 Alu, 5 L1, and 7 SVA for GMKF and 2, 4, 2 for 1KGP, respectively; Fig. 1c). The prevalence of mosaic DNRTs in 1KGP (17%) could be explained by somatic DNTRs occurring in cell culture. Indeed, a higher mosaic rate for SNP/Indel has been reported on the same data, attributed to ongoing mutation processes in cell culture (Byrska-Bishop et al. 2022; Ng et al. 2021). As a comparison, the mosaic DNRTs in GMKF was 12%, likely due to early embryonic events. This result is consistent with an independent study on normal colon that identified one mosaic L1 insertion occurring at the fourth cell division (Nam et al. 2022).

For the 142 (out of 162) and 38 (out of 46) germline DNRTs in GMKF and 1KGP respectively, we ran trio-based phasing using nearby heterozygous SNPs (see Method for details) to determine their parental origins. Limited by the read length and insert size of the sequencing libraries, we were only able to phase only 26 out of 142 GMKF DNRTs and 13 out of 38 1KGP DNRTs (Fig. 3d). Nonetheless, our inspection of the phased DNRTs by repeat type revealed a significant enrichment (p=0.019; binomial test) for *Alu* DNRTs of paternal origin in the GMKF cohort while no statistical significance was reached for L1 and SVA (Fig. 3e). A similar trend was also observed in the 1KGP cohort, although it was not statistic significant due to small sample size (Fig. 3e). A similar pattern was also reported in an earlier study on large pedigree data of three generations (Feusier et al. 2019). Based on these results, we hypothesize that the *Alu* insertion rate is higher in the cell types involved in spermatogenesis compared to oocytes.

### *De novo* rate estimation and enrichment of deleterious retroelements

The large number of trios allowed us to estimate the *de novo* rate of the retroelements in the birth defect cohorts. Previous estimates were highly variable (Borges-Monroy et al. 2021), with our recent study on the Simons Simplex Collection (SSC) of ASD cases (2,288 families with proband and siblings) using an earlier version of xTea giving 1/26 birth (adjusted to 1/21 after considering detection sensitivity benchmarked from long reads). Below, we computed the exact binomial confidence intervals on all the GMKF cohorts combined as well as for four disease groups that have sufficient sample sizes: Congenital Heart Defect (CHD), Congenital Diaphragmatic Hernia (CDH), Orofacial Defect, and Pediatric Tumor. The Orofacial Defect group consists of 3 cohorts (phs001997, phs001420, and phs001168 in Fig. 1b) of the same defect but from different populations. The Pediatric Tumor group consists of 4 cohorts (phs001683, phs001228, phs001846, and phs001436 in Fig. 1b) of different tumor types. For reference, we list the *de novo* insertions for two earlier studies on the SSC ASD (Tab. S3), 1KG (Tab. S4), and the Utah Centre d’Etude du Polymorphisme Humain (CEPH) cohorts (Tab. S5).

The *de novo* retroelement rate by repeat type are shown in Fig. 4a for the different cohorts. The rate of all the GMKF cohorts is around 1/34 (1 per 34 births) for *Alu*, 1/108 for L1, and 1/93 for SVA, and 1/20 for all 3 TE types combined. Compared to the ASD cohort, we observed a similar rate (1/34 vs. 1/35) for *Alu* insertions, but much higher rate (1/108 vs. 1/189) for L1 and (1/93 vs. 1/244) SVA insertions. Our results are consistent with a previous study (Deciphering Developmental Disorders Study 2017) that showed a higher rate for *de novo* SNVs/indels in the DDD cohort compared to the ASD cohort.

**Fig. 4:**
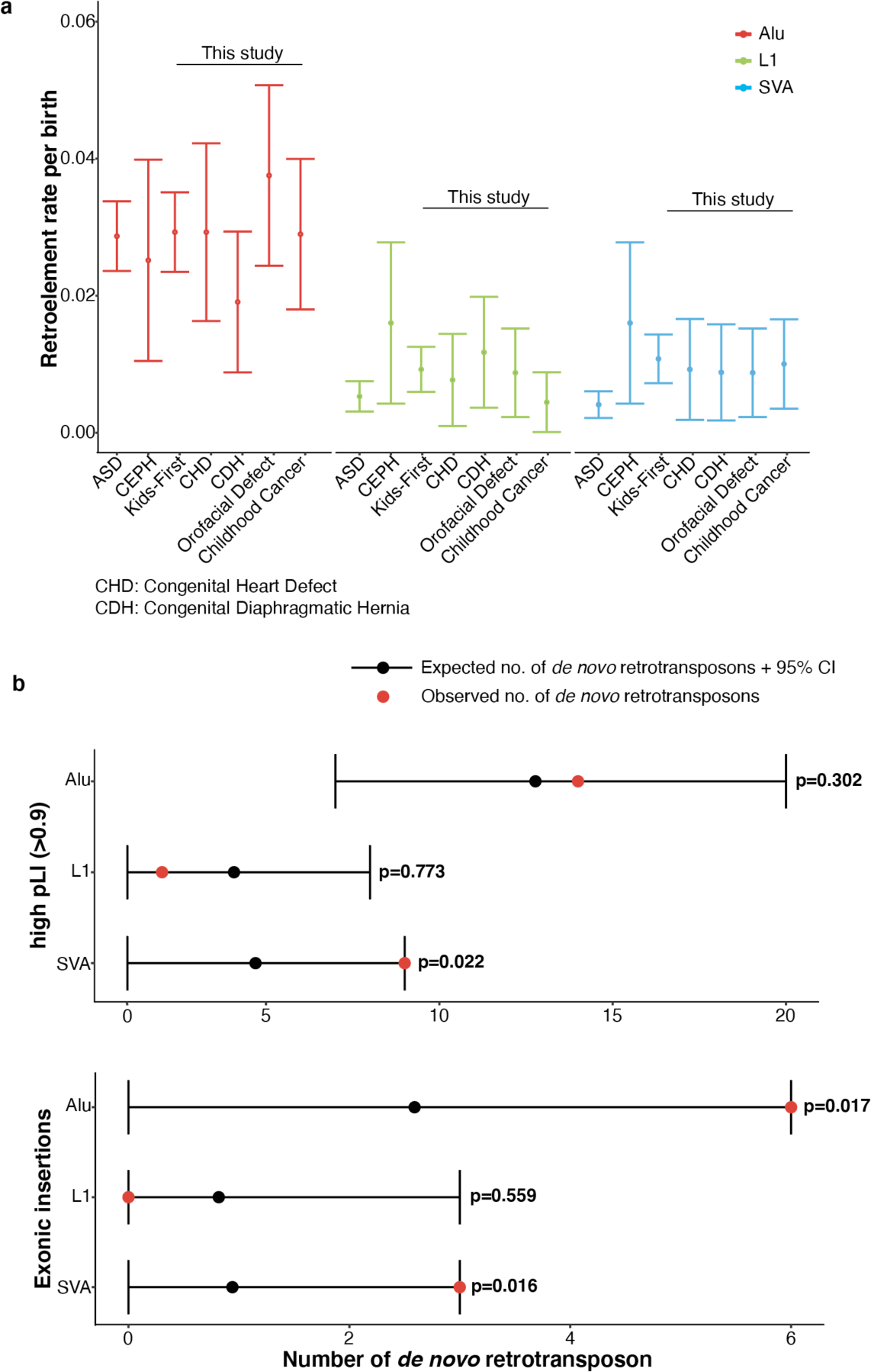
*De novo* rate and enrichment analysis of retroelements. **a** We calculated the *de novo* rate and 95% confidence interval using an exact binomial confidence interval estimate with x=number of retroelements and N=number of births. ASD and CEPH are from two previous studies and the rest are from the GMKF cohorts in this study. GMKF is for all the 12 cohorts, “Orofacial Defect” is combined from 3 orofacial defect cohorts (phs001997, phs001420, and phs001168), and “Childhood Cancer” is combined from 4 cancer related cohorts (phs001683, phs001228, phs001846, and phs001436). The *de novo* rate for L1 and SVA in this study is clearly higher than ASD, with a similar rate for *Alu*. **b** We checked whether the identified de novo retroelements from the GMKF cohorts were enriched in genes whose pLI>0.9 (top) or enriched in exonic regions (bottom). We ran 10,000 simulations (details in Method) for *Alu*, L1 and SVA, and compared with the number of observed ones. There is a statistically significant enrichment of SVA insertions fallen in genes with pLI>0.9. Both *Alu* and SVA insertions were also found enriched in exonic regions.

Given the higher rate of insertions in the GMKF cohorts, we examined whether the DNRTs are enriched in genes likely to be intolerant of loss of function (LoF), as determined by the pLI score (Lek et al. 2016) and whether they enriched in exonic regions. We first simulated the distribution of DNRTs across the genome with the following idea. When L1 insertions (also for *Alu* and SVA as they rely on the L1 protein) are integrated into the genome, they prefer specific motifs (consensus TTTTT/AA). Inspired by the landmark experimental study on L1 endonuclease activity (Flasch et al. 2019) that engineered L1 insertions in cultured cell lines to characterize the pattern of the cleavage sites, we simulated the insertion sites based on the frequency of cleavage motif sequences. A key step here is to construct the weight matrix based on the frequency of the cleavage motifs. We calculated the frequency from germline insertions, different from the experimental study (Flasch et al. 2019) that inferred the matrix from engineered somatic insertions, but the logo plots of cleavage site sequences were almost identical (Fig. S2; see Method for details on the simulation procedure). Compared to the background distribution generated from 10,000 simulations, we observed an enrichment (p=0.022) for *de novo* SVA insertions in the high pLI (>0.9) genes, but not for *Alu* and L1. We also find an enrichment of *de novo Alu* (p=0.017) and SVA (p=0.016) insertions in the exons, but no statistical significance for *de novo* L1 insertions (Fig. 4b). Both analyses suggest that *de novo* retrotransposons—especially *de novo* SVA insertions—found in the birth defect cohorts are more likely to be deleterious.

### *De novo* processed pseudogene insertion and *de novo* TE insertion activity

Although germline and somatic PPG insertions in human have been reported (Esnault et al. 2000; Feng and Li 2021; Schrider et al. 2013; Ewing et al. 2013; Cooke et al. 2014), *de novo* PPG insertions have not been well characterized due to their low *de novo* rate and the lack of detection tools designed for PPGs. The study on the DDD cohort reported 2 *de novo* PPG insertions from 9,738 WES trios (Gardner et al. 2019). Here, we extended our xTea method for *de novo* PPG insertion detection and identified 2 *de novo* PPG insertions from 3,244 trios (Fig. S4), suggestive of a 3-fold increase in insertion rates. One insertion originated from gene *HNRNPM* and reverse-transcribed into a truncated PPG insertion within the intronic region of *FBXL7*. The other originated from gene *ZNF664* and mobilized to an intergenic region on chromosome 2 (Fig. 5a; S4).

**Fig. 5:**
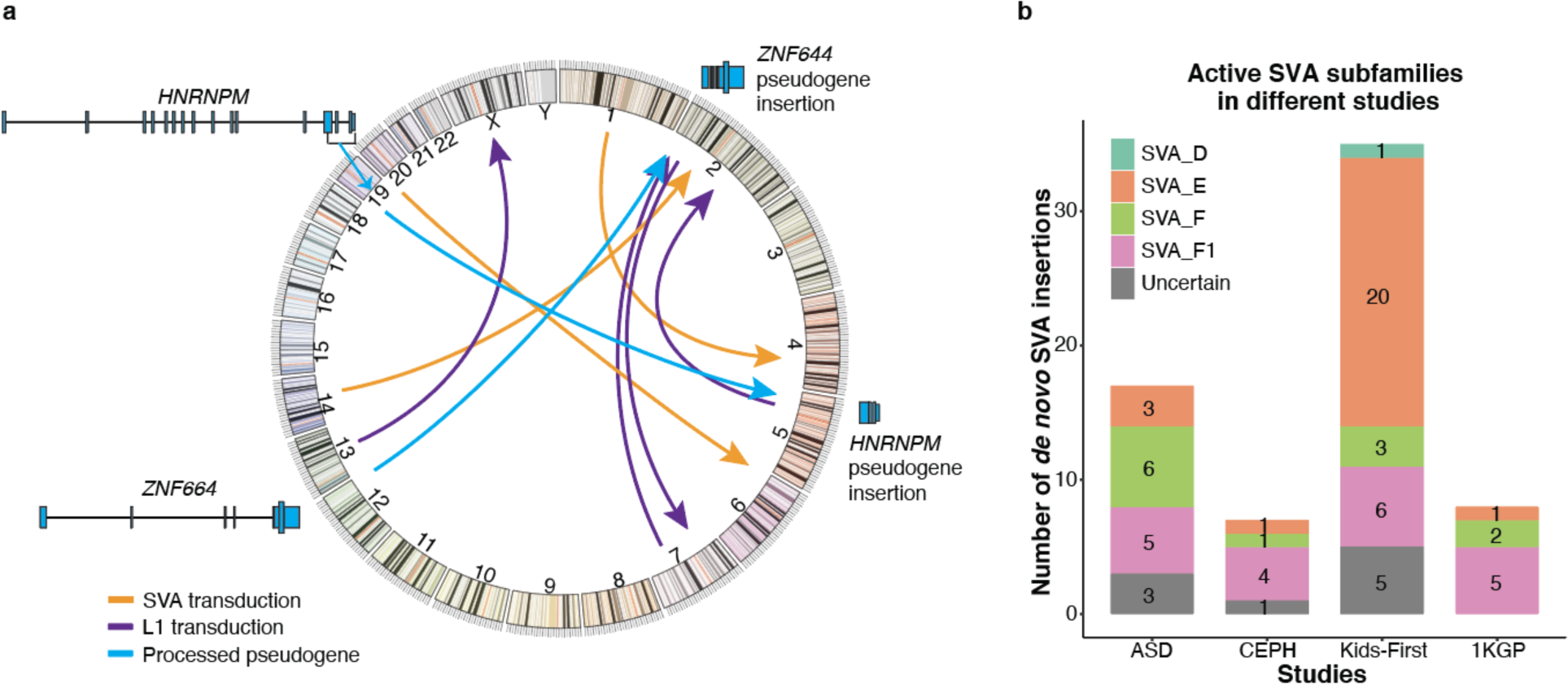
Activity of *de novo* retroelements. **a** 4 L1 transductions, 3 SVA transductions, and 2 pseudogene insertions were identified. Each arrow points from the source elements to the insertion site. Specifically, the 2 pseudogene insertions were originated from gene *HNRNPM* and gene *ZNF664*, with *HNRNPM* insertion is truncated while the *ZNF664* one is of full length. **b** For all the *de novo* SVA retrotransposon insertions identified from the four studies (17 ASD, 7 CEPH, 35 GMKF and 8 1KGP), we further checked the subfamilies of each insertion. 3, 1, 5 were not well annotated from ASD, CEPH, and GMKF respectively. Out of those well annotated ones, SVA_F1 (20/58) and SVA_E (25/58) are the most active subfamilies.

For L1 and SVA, the flanking regions (mainly 3’ for L1, and 5’ and 3’ for SVA) sometimes transpose together with the retrotransposon to form transductions (TDs). Because most of the TD sequences are unique in the genome, they could be used to trace the source elements of the insertions. We ran xTea on all the proband cases having *de novo* L1 and SVA insertions and identified 4, 1, 1, and 1 transduction from GMKF, 1KGP, ASD and CEPH cohort, respectively. In the original CEPH paper (Feusier et al. 2019), the authors reported 3 L1 TDs, but after manual inspection, we could only confirm one of them. Our results indicate a small number of active *de novo* transduction events.

Finally, we investigated the activity of *de novo* insertions for SVA, one of the youngest retrotransposons in the human genome. SVA_E and SVA_F subfamilies are the known major active subfamilies (Wang et al. 2005; Hancks and Kazazian 2010); SVA_F has two subfamilies, SVA_F1 and CH10_SVA_F, that fused with part of *MAST2* to form new structure and transpose. CH10_SVA_F has two *Alu* copies at the two ends, making the annotation from short reads difficult; thus, the activity of CH10_SVA_F is not explored here. SVA_F1 insertions can have reads on one side that cover the *MAST2* region and reads on the other sides aligned to the SVA_F region, thus allowing for annotation. Using an SVA annotation module we recently developed (Chu et al. 2023), we examined each of the *de novo* SVA insertions from the ASD (n=17), CEPH (n=7), GMKF (n=35), and 1KGP (n=8) study (Fig. 5b) and found that SVA_E (25/58; 9 could not be classified) and SVA_F1 (20/58) are the two most active SVA subfamilies in human genome.

## DISCUSSION

With a systematic analysis of the 12 GMKF WGS cohorts, our results revealed the different pathogenic roles DNRTs may have in causing birth defects and childhood cancers. Our study highlights the importance of WGS in identifying causal mutations beyond the standard variant types and the effectiveness of the improved xTea pipeline we developed for characterizing DNRTs in large cohort analyses and disease diagnosis.

Compared to germline DNRTs, mosaic DNRTs occurring in the proband are substantially more difficult to identify due to their low VAFs. Our analysis found a set of DNRTs with low VAFs, with a peak around 25% in the VAF density plot for GMKF. Although they could be variations in read sampling or germline DNRTs in aneuploid regions, we suspect that at least some of them are mosaic variants that occurred very early in embryonic development. A peak at the similar VAF (∼ 20%) was observed for 1KGP but those are likely to be somatic variants that arose in cell culture; for ASD cohorts, no such peaks were found, increasing the likelihood that the peak in the GMKF cohort may be due to mosaic variants. A further analysis on other types of mutations (especially *de novo* SNP/Indels) or sequencing data with higher coverage may provide additional information for the prevalence of mosaic mutations.

Strong support for the functional importance of *de novo* SVA insertions came from their enrichment in genes that are estimated to be intolerant to loss of function (pLI>0.9) as well as exonic enrichment for *de novo* SVA and *Alu* insertions. We also identified several cases in which DNRTs disrupted genes that are associated with the disease in the literature. Most notable are the *Alu*-promoted deletion and an exonic insertion on the *NF1* gene. Together with the earlier studies (Wallace et al. 1991; Vogt et al. 2014; Wimmer et al. 2011) on retroelements on *NF1* gene, our study indicates a hot spot for RTs on the *NF1* gene. For other cases, experimental validation is needed to ascertain whether those candidate DNRTs are causative of the observed phenotypes.

Most studies on retroelement insertions have focused on germline L1 and *Alu* insertions, as they are the easiest to identify due to their relatively simple structure. SVA are substantially more complicated in its structure, especially with the VNTR (variable number of tandem repeats) region in the middle often causing mis-annotation, as revealed in our recent work on SVA detection and annotation (Chu et al. 2023). The increasing adaption of long-read sequencing platforms will greatly enhance detection sensitivity and specificity, as well as expanding the regions of the genome that can be interrogated. The GMKF cohort continues to increase in size, as do numerous other WGS studies for various genetic diseases. The resulting datasets will enable discovery of additional disease-associated genes, allow for more accurate inferences on the rates of mosaic insertion events, help pinpoint active source elements through transduction events, and shed light on the expanding role that retroelements play in disease initiation.

## METHOD

### *De novo* retroelements identification from trio data

Compared to germline retroelement insertions, DNRTs are rare, thus sensitivity is important for identification. Here, we optimized our xTea method for DNRT insertion identification. Fig. 1a shows the major steps: for one given trio data, *(i)* we ran xTea germline module on the proband sample (by default with parameters “--nclip 2 --cr 0 --nd 3 --nfclip 2 --nfdisc 3”); *(ii)* we ran xTea somatic module on the candidates generated from step *(i)* with alignments from both parents as controls; *(iii)* we further developed a machine learning based filtering module (manuscript in preparation) to filter out the false positives. For each candidate insertion, we converted the alignments to images using BamSnap (Kwon et al. 2021), where each candidate is composed of three images from the trio on the same location. We first prepared a positive training set from semi-simulated data, where we selected germline heterozygous retroelements from one sample and viewed it as the “proband”, and then we selected two unrelated samples that do not have retroelement insertions on this location as the “parents”. From these three “combined” samples on this location we generated one positive image. In this way, we prepared 6952 positive training images. Then we prepared the same number of negative training images from xTea output on two cohorts (phs001228 and phs001168) that we had manually inspected for the true positives. Next, we trained a model from the positive and negative training sets. Then, for each candidate image, we predict it to true positive or false positive; *(iv)* lastly, we ran manual inspection on each of the candidates to select the true positive variants.

### *De novo* retroelements annotation

For each identified DNRT, we first annotated it as exonic, 5’ UTR, 3’ UTR, intronic, or intergenic based on the GENCODE (v28 on GRCh38) gene annotation file. Then, we ran ANNOVAR (version downloaded on May 2022) to annotate DNRTs fall in promoter and enhancer regions. We used the pLI score from the ExAC study (Lek et al. 2016) to annotate the estimated intolerance of each gene to mutations, and pLI>0.9 are annotated as “high intolerant”.

In addition, we also ran subfamily annotation specifically for *de novo* SVA insertions, because the active subfamilies are less well characterized in large cohorts. To annotate the SVA insertion subfamilies, we first collected all the discordant and clipped reads originated from the insertion. Then we ran local assembly on the collected reads using the xTea assembly module. Next, we ran the SVA annotation module (initially developed for a different study whose manuscript in review) on the assembled sequences to get the subfamily information of each *de novo* SVA insertion. Insertions annotated to more than one subfamily or failed to be assembled were annotated as “uncertain”.

### Variation allele frequency (VAF) calculation

Different from SNV VAF estimated by calculating the ratio between the number of reads containing the mutation and the total number of reads at the site, calculating VAF for DNRTs is more challenging. We illustrate the major biases introduced due to reads alignments in Fig. S1. Generally, for one clipped read with part of the read from the retroelements and the other part from the flanking regions, if the length of the retroelement part is short (by default, BWA mem has a minimum kmer length of 19), then the read will be aligned elsewhere as the unique part (part from flanking region) is short, as a result, these reads will not be counted when calculating the VAF. To correct the introduced bias, when counting the number of full mapped reads covering the breakpoint, we skipped reads having short overlap (by default <19) with the flanking region. In addition to calculate VAF from reads covering the breakpoint, discordant pairs can also be used for calculating the VAF. Basically, within the given range (by default, insert-size) we count the number of discordant pairs and concordant pairs, and then calculate the VAF. In Fig. S1, we show the calculated clip- and discordant-based VAFs for 115,115 germline TE insertions. In practice, we took the average of the clip- and discordant-based VAF as the VAF of each DNRT.

### Trio based phasing

For some of the identified DNRTs, we can phase them to derive paternal or maternal origin based on the nearby heterozygous SNVs. For example, if we find heterozygous SNVs in the father and identify the same SNVs within the discordant pairs of the DNRT in the proband, then we can infer the DNRT to be of paternal origin (Fig. S3c,d). However, in practice, we can only find nearby heterozygous SNVs for a small fraction of DNRTs. To broaden the range of phasable DNRTs, for the germline DNRTs in proband, if we observe heterozygous SNVs in one of the parents, but the same SNVs are found in the non-DNRT reads, then we can infer that the DNRT is inherited from the other parent. To achieve this, we first adopt a Gaussian Mixture Model (2 mixture components) to classify the DNRTs as germline or mosaic, and then phase the set of germline variants. To call heterozygous SNVs from the parents, we ran samtools “mpileup” and “call” on the local region of each DNRT. To call SNVs for each DNRT from the proband, we first separated the reads aligned to the local region to two groups: “DNRT reads” and “non-DNRT reads”. For each group, we ran samtools “mpileup” and “call” to identify the SNVs. In addition, we also ran manual inspection for each phased DNRTs from BamSnap screenshots to further validate the phased the DNRTs. The whole pipeline is shown in Fig. S3.

### *De novo* rate estimation

To estimate the *de novo* rate of retroelements, we adopted the exact binomial confidence interval estimate, where X is the number of retroelements, and N is the number of births. To compare the *de novo* rate among different disease cohorts, we show the results of “Syndromic cranial dysinnervation”, “Heart birth defects”, “Orofacial birth defects”, “childhood cancer”, where “Orofacial birth defects” results are merged from three orofacial birth defects of different populations (phs001168, phs001420, and phs001997), and the “childhood cancer” results are merged from the four tumor cohorts (phs001436, phs001228, phs001683, and phs001846). In addition, we also compared the overall *de novo* rate in the birth defect disorders with two earlier studies on autism (Borges-Monroy et al. 2021) and large pedigree of normal samples (Feusier et al. 2019), where we used the number of *de novo* retroelements and the number of births in their released results.

### *De novo* retroelements enrichment analysis

To test whether the identified DNRTs are enriched in the GMKF data or not, we need a control model that simulates the random hits of DNRTs. However, it is known that RTs are not purely randomly happened on the genome. A recent study based on engineered L1s in cell lines inferred that there are specific endonuclease (EN) cleavage motifs (Flasch et al. 2019). Here, we adopt the similar approach to build the control model as described in the endonuclease activity study from engineered L1s (Flasch et al. 2019). As shown in Fig. S2, we first gather all the possible EN cleavage motifs, then for each motif we estimate its frequency, which later will be used as the probability of a simulated insertion occurring with the motif. Differently, here we use germline insertion rather than engineered *de novo* L1s to gather all the possible motifs. To achieve this, we first ran xTea on the 1KGP high depth WGSs and collect the high-quality TE insertions (labeled with “tprt_both” in xTea output indicating exist of both the TSD and polyA tail; and require the population AF>0.01). For each TE insertion, we collect the first left 4 bases and the right 3 bases at the breakpoint (adjusted accordingly for antisense cases). Then, we put all the collected motifs in a table and calculate the frequency for each one. We also adopt the same approach as described in the mentioned study (Flasch et al. 2019) to include potential motif not recruited in the motif table: We split the 7-base motif to 3 independent segments: first 2 bases, middle 4 bases and the last base. For each segment, we calculate the frequency of each sub-motif based on the frequency of the 7-base motifs. In this way, we have 3 tables of sub-motifs whose frequency have been calculated. Thus, to generate a 7-base motif, we generate the 3 segments separately and for each segment we select a sub-motif based on the frequency table. Then, we merge the 3 segments to one 7-base motif. Now, given one motif, we need to find out where on the genome this motif can be generated. To achieve this, we build another 7-base motif table for the whole genome, where we save all the positions of each motif. In this way, once given a motif, we randomly select one from all the positions.

To construct the control model, for each round we simulated the same 163 DNRTs with our pipeline, and we repeated the experiments for 10,000 times. We did enrichment analysis for insertions fallen in exon regions and in pLI high (>0.9) genes separately.

## Data availability

The 12 cohorts of pediatric whole genome sequencing (WGS) data were accessed through the portal of Gabriella Miller Kids First Pediatric Research Program https://portal.kidsfirstdrc.org/. The high depth trio based WGS data from the 1000 Genomes Project were downloaded from the International Genome Sample Resource (IGSR) at https://www.internationalgenome.org/data/.

## Code availability

Source code for the de novo retroelements identification is available at https://github.com/parklab/xTea (doi:10.5281/zenodo.4743788).

## Acknowledgements

This work was supported by R03CA249364 from the National Cancer Institute. VL is supported by the Swedish Research Council (2020-00583).

